# Increased bacterial taxonomic and functional diversity is associated with impaired rotavirus vaccine immunogenicity in infants from India and Malawi

**DOI:** 10.1101/2023.03.24.23287614

**Authors:** Edward Cunningham-Oakes, Christina Bronowski, End Chinyama, Khuzwayo C. Jere, Kulandaipalayam Natarajan C. Sindhu, Gagandeep Kang, Miren Iturriza-Gómara, Alistair C. Darby, Edward P. K. Parker, the RoVI study group

## Abstract

The immunogenicity and effectiveness of oral rotavirus vaccines (ORVs) against severe rotavirus-associated gastroenteritis is impaired in low- and middle-income countries (LMICs) where the burden of disease is highest. Determining risk factors for impaired ORV response may help identify strategies to enhance vaccine effectiveness. In this study, we use metagenomic sequencing to provide a high-resolution taxonomic analysis of stool samples collected at 6 weeks of age (coinciding with the first ORV dose) during a prospective study of ORV immunogenicity in India and Malawi. We then analyse the functional capacity of the developing microbiome in these cohorts. Microbiome composition differed significantly between countries, although functional capacity was more similar than taxonomic composition. Our results confirm previously reported findings that the developing microbiome is more diverse in taxonomic composition in ORV non-seroconverters compared with seroconverters, and we additionally demonstrate a similar pattern in functional capacity. Although taxonomic or functional feature abundances are poor predictors of ORV response, we show that skews in the direction of associations within these microbiome data can be used to identify consistent markers of ORV response across LMIC infant cohorts. We also highlight the systemic under-representation of reference genes from LMICs that limit functional annotation in our study (7% and 13% annotation at pathway and enzyme commission level, respectively). Overall, higher microbiome diversity in early life may act as marker for impaired ORV response in India and Malawi, whilst a holistic perspective of functional capacity may be hidden in the “dark matter” of the microbiome.

## INTRODUCTION

The global roll-out of oral rotavirus vaccines (ORVs) has formed a cornerstone of public health efforts to mitigate the morbidity and mortality associated with infant diarrhoeal disease. Since the licensure of RotaTeq and Rotarix in 2006 and 2008, respectively, ORVs have been introduced into the national immunisation programme of over 100 countries. In conjunction with improvements in sanitation infrastructure, ORV implementation has helped bring about a reduction in rotavirus-associated mortality from over 500,000 in the year 2000 to approximately 130,000 in 2016^1^. However, the potential impact of ORV is constrained by the impaired effectiveness of available vaccines in low- and middle-income countries (LMICs), where the vast majority of rotavirus-associated deaths occur^2,3^.

Much uncertainty remains regarding the mechanisms underpinning the ORV effectiveness gap. Early microbial exposures play a key role in shaping immune system development. Accordingly, several recent studies have applied high-throughput 16S ribosomal RNA (rRNA) gene sequencing to explore the extent to which infant gut microbiota development might influence ORV response. While some have yielded significant associations^4^, others have not^5^, and consistent microbial signatures associated with ORV response remain elusive. In one of the largest studies of this phenomenon to date, we applied standardised methods to explore the link between microbiota development (assessed via 16S rRNA sequencing in multiple longitudinal samples) and ORV response across cohorts in India, Malawi, and the UK^6^. Notably, while we did not identify clear associations between taxonomic abundances and ORV outcome, microbiota diversity at 6 weeks of age (the timing of the first dose of ORV) was negatively correlated with rotavirus seroconversion status in both Indian and Malawian infants.

Compared with 16S-based methods, metagenomic sequencing has the potential to offer more granular insight into the taxonomic composition and functional capacity of the developing microbiome. Herein, we applied standardised metagenomic sequencing methods to stool samples collected at 6 weeks of age in India and Malawi. Our study highlights a more diverse functional and taxonomic profile in ORV non-responders compared with responders in both countries, confirming and extending our previous findings based on 16S sequencing^6^.

## METHODS

### Study population

The Rotavirus Vaccine Immunogenicity (RoVI) study was a multi-site observational cohort study that explored the impact of maternal antibodies, microbiota development, and environmental enteric dysfunction (EED) on infant ORV response (CTRI/2015/11/006354). Details regarding study design, sample handling, and primary outcomes of the RoVI study have been described previously^6,7^. Pregnant women were enrolled in Blantyre (Malawi; n = 187), Vellore (India; n = 395), and Liverpool (UK; n = 82). Routine immunisations were administered according to the national immunisation schedule at each study site, including two doses of Rotarix at weeks of life 6 and 10 (India and Malawi) or weeks of life 8 and 12 (UK). Recruitment (by month of birth) spanned December 2015 to November 2016 in India and November 2016 to April 2018 in Malawi. Rotavirus-specific IgA (RV-IgA) was measured in infant blood samples collected pre-vaccination (before dose 1) and post-vaccination (4 weeks after dose 2). Seroconversion was defined as detection of RV-IgA at ≥20CIU/ml post-vaccination among infants who were seronegative at baseline or a 4-fold increase in RV-IgA concentration among infants who were seropositive at baseline. Rotavirus shedding was assessed in six stool samples per infant, including measurement of the Rotarix *NSP2* gene in samples collected 1 week after each ORV dose.

### Sample processing and sequencing

DNA extraction from stool was carried out using the QIAamp DNA stool mini kit (Qiagen) according to the manufacturer’s instructions, with a number of modifications as described previously^6^. Extracted DNA was stored at -70°C until library preparation. Overall, 16S rRNA gene sequencing was carried out for a total of 2,137 separate stool samples across the three cohorts. For the metagenomic follow-up study, we focused on stool samples collected at 6 weeks of age in India and Malawi given the significant negative correlation between microbiota diversity and rotavirus seroconversion that was evident at this time point in both cohorts. Stool samples were selected based on the following eligibility criteria: (i) the sample yielded a high-quality 16S rRNA microbiota profile (≥25,000 sequences after quality filtering); (ii) the infant met the primary study endpoint (measurement of seroconversion or dose 1 shedding); and (iii) sufficient volume of extracted DNA (>1 μl) was available to proceed with metagenomic library preparation. A total of 355 samples fulfilled these eligibility criteria (288 and 67 from India and Malawi, respectively). Library preparation and sequencing was performed at the Centre for Genomic Research (University of Liverpool). Sample order was randomised. Libraries were prepared using the NEBNext^®^ Ultra^™^ II FS DNA Library Prep Kit for Illumina and sequenced via two lanes of 2 × 150bp paired-end sequencing on an Illumina NovaSeq 6000 instrument with S4 chemistry (with all samples included in both lanes).

### Pre-processing of reads

Adapter sequences were removed from reads using Cutadapt^8^ (version 1.2.1), trimming the 3’ end of any reads which match the adapter sequence for 3 bp or more. Sickle (version 1.200) was then used for further trimming of reads, with a minimum window quality score of 20. Any reads shorter than 15 bp after trimming were removed. Human DNA was removed from all paired-end reads using the MetaWRAP^9^ read_qc module, which makes use of BMTagger^10^ to remove human contamination, via alignment to the hg38 reference genome (obtained from ftp://hgdownload.soe.ucsc.edu/goldenPath/hg38/chromosomes/).

### Taxonomic assignment

Taxonomy was assigned to reads by Kraken2^11^ (version 2.1.2), using a custom database that included RefSeq complete genomes and proteins for archaea, bacteria, fungi, viruses, plants, and protozoa, as well as RefSeq complete plasmid nucleotide and protein sequences, and a subset of the NCBI UniVec database. A confidence threshold of 0.1 was set for read assignments, and reports were generated for downstream biom file generation. Read counts assigned to taxonomies in each sample were then re-estimated using the average read length of that sample, using Bracken^12^ (version 2.0). Kraken-biom (version 1.0.1) was then used to generate biom file in json format, using Bracken reports. Biom (version 2.1.6) was then used to assign tabulated metadata to this biom file. Reflecting the filtering steps applied to the 16S sequencing data, species were retained for the analysis if they were bacterial and detectable at an abundance of ≥0.1% in at least two samples.

### Functional assignment

Gene pathway and enzyme commissions (ECs) were assigned to reads using HUMAnN^13^ (version 3.0.0) with default parameters. Pathway assignments were produced as a default output of HUMAnN, whilst ECs were assigned using the utility scripts *humann_rename_table* and *humann_regroup_table* to rename gene family table features. Reads that were unmapped (mapped to gene families without UniRef90 identifiers), unintegrated (reads that were assigned to a gene family but could not be grouped into pathways), or ungrouped (reads that were assigned to a gene family but could not be assigned to an EC) were retained in the analysis. For each analysis level (pathway and EC), unstratified outputs were merged into a feature table and renormalised to copies per million (cpm) within HUMAnN. Renormalised feature tables served as input data for downstream statistical analysis.

### Statistical analysis

All analyses were performed in the programming language R (version 4.2.1). To assess the consistency of 16S versus metagenomic sequencing, we compared genus-level relative abundances and richness estimates between the pipeline outputs via Spearman’s rank correlation coefficient (rho) with two-sided hypothesis testing. These comparisons were performed at a rarefaction depth of 25,000 reads per sample for 16S data (as per our prior analyses of these data) and 1 million reads per sample for metagenomic data. All subsequent analyses were performed in parallel for four modules of input data: (i) metagenomic taxonomy based on species-level relative abundances; (ii) metagenomic taxonomy based on genus-level relative abundances; (iii) metagenomic relative pathway abundances; and (iv) metagenomic relative EC abundances. Genus- and species-level relative abundances were determined at a rarefaction depth of 1 million reads per sample. Pathway and EC relative abundances were calculated based on renormalised counts in cpm. Primary outcomes of interest included country, seroconversion status, dose 1 shedding status, and post-vaccination RV-IgA concentration. For ORV outcomes, we performed separate analyses for Indian and Malawian infants to account for significant geographic differences in baseline microbiome composition. For Indian infants, we also stratified analyses by neonatal rotavirus infection status – defined as the detection of wild-type rotavirus shedding in week of life 1 and/or baseline seropositivity (pre-vaccination RV-IgA ≥20 IU/ml) – given the impact of this exposure on microbiota-related associations in our prior analyses^6,7^.

Differences in alpha diversity (feature richness at species, genus, pathway, or EC level) were assessed via Wilcoxon rank-sum test for binary outcomes and Spearman’s rank correlation coefficient for RV-IgA. Differences in beta diversity were assessed based on unweighted Bray–Curtis distances via permutational analysis of variance (PERMANOVA) with 999 permutations. Multivariate linear-regression analysis via MaAsLin2^14^ (version 1.8.0) was used to identify differentially abundant features for each outcome of interest. For ORV outcomes, we adjusted for the following covariates: age at sample collection (in days); antibiotic exposure (any exposure up to 14 weeks of age); birthweight (in kg); breastfeeding status (exclusive, partial, or none up to 11 weeks of age); and delivery mode (caesarean or vaginal; only included in India given that all births in Malawi were via vaginal delivery). We also compared feature prevalence (presence/absence) in relation to each outcome of interest via Fisher’s exact test (for binary outcomes) and Wilcoxon rank-sum test (comparing RV-IgA in infants with versus without the feature of interest, excluding features with a prevalence of <5% or >95%). *P* values for differential abundance testing were adjusted by Benjamini–Hochberg false discovery rate (FDR) correction and a significance threshold of <0.2 applied for the purposes of this exploratory analysis. For adjusted MaAsLin2 outputs, covariate-associated P values were excluded prior to FDR correction to ensure that the adjusted *P* value distribution related specifically to the outcome of interest. As a complement to differential feature selection via FDR *P* value threshold, we assessed potential skew in the associations for each outcome of interest using Wilcoxon’s rank-sum test (with the null hypothesis that positive and negative associations occur at equal frequency). We also explored the strongest associations (top 10% per data module) with ORV outcome to determine consistency of selected features across countries.

We applied the Random Forests algorithm to predict country, seroconversion, or shedding status (classification approach), or post-vaccination log-transformed RV-IgA (regression approach) based on feature abundances, as described previously^6^. For each model, 20 iterations of 5-fold cross-validation were performed. Baseline accuracy in classification models was standardised at 50% by fitting each iteration of 5-fold cross-validation on a random subset of 50 samples per group (or the number of samples in the minority group if this was <50). For regression models, out-of-bag R^2^ values for predicted vs observed RV-IgA values were determined via linear regression. Variable importance was determined based on Gini index (classification) or mean squared error (regression). Features were included in differential abundance analyses and Random Forests models if they were detected with a prevalence of >5% in at least one analysis group.

To supplement the primary outcome analyses, we performed an exploratory analysis of demographic covariates associated with metagenome composition and function. We report on variables associated with alpha diversity (assessed via Wilcoxon rank-sum test for binary covariates and Spearman’s rho for linear covariates) or beta diversity (assessed via PERMANOVA of unweighted Bray-Curtis distances), applying a significance threshold of <0.2 after FDR adjustment.

## RESULTS

A total of 341 samples (283 and 58 from India and Malawi, respectively) yielded high-quality metagenomic taxonomy profiles (≥1 million sequences after quality filtering) and were retained in the final analysis. Baseline characteristics and ORV outcomes are summarised for each cohort in **Table 1**. Indian infants were more likely than Malawian infants to be born by caesarean delivery and exhibited high rates of neonatal wild-type rotavirus infection, while Malawian infants were more likely to be HIV exposed. Exclusive breastfeeding predominated in both cohorts, and antibiotic exposure across the study period was reported at a similar rate (72% vs 74% in India vs Malawi).

**Table 1.** Baseline characteristics of study cohorts.

|  | India | Malawi |
| --- | --- | --- |
| *N* | 283 | 58 |
| Caesarean delivery | 62/283 (21.9) | 0 (0.0) |
| Birthweight (kg), *mean (s.d.)* | 2.96 (0.43) | 3.02 (0.45) |
| Female |  |  |
| Polio vaccine schedule |  |  |
| Trivalent OPV | 51 (18.0) | — |
| Mixed trivalent/bivalent OPV* | 56 (19.8) | — |
| Bivalent OPV* | 84 (29.7) | 58 (100.0) |
| IPV§ | 92 (32.5) | — |
| Exclusive breastfeeding up to 11 weeks of life | 243 (85.9) | 53 (91.4) |
| Exposed to antibiotics up to 14 weeks of life | 204 (72.1) | 43 (74.1) |
| HIV status |  |  |
| Exposed | 0 (0.0) | 15 (25.9) |
| Unexposed | 275 (97.2) | 41 (70.7) |
| Unknown | 8 (2.8) | 0 (0.0) |
| Neonatal rotavirus infection | 149/281<sup>†</sup> (53.0) | 6/47<sup>†</sup> (12.8) |
| Seroconverted to ORV | 78/282<sup>†</sup> (27.7) | 10/56<sup>†</sup> (17.9) |
| Shed ORV at week of life 7 (1 week after dose 1) | 74/281<sup>†</sup> (26.3) | 21/47<sup>†</sup> (44.7) |
| RV-IgA at week of life 14 (4 weeks after dose 2), *GM (95% CI)* | 18.9 (14.9–23.9) | 6.4 (4.0–10.2) |

Data are n (%) unless otherwise stated. Seroconversion was defined as detection of RV-IgA at $\geq 20$ IU/ml post-vaccination among infants who were seronegative at baseline or a 4-fold increase in RV-IgA concentration among infants who were seropositive at baseline. Neonatal rotavirus infection was defined as the detection of wild-type rotavirus shedding in week of life 1 and/or baseline seropositivity (pre-vaccination RV-IgA $\geq 20$ IU/ml). \*Enrolment spanned the global transition from trivalent to bivalent OPV in April 2016. § A sequentially recruited cohort in India received IPV instead of OPV at 6 and 10 weeks of age; <sup>†</sup> Infants with missing data excluded. GM, geometric mean; IPV, inactivated poliovirus vaccine; OPV, oral poliovirus vaccine; ORV, oral rotavirus vaccine; RV, rotavirus.

### Metagenomic taxonomic profiles closely matched 16S profiles

Genus-level richness estimates generated by 16S and metagenomic sequencing were strongly correlated (Spearman’s rho = 0.80; **Supplementary Figure 1A**), as were the relative abundances of the 10 most abundant genera (rho >0.8 for 8 of 10; **Supplementary Figure 1B**). However, the genus *Bifidobacterium* tended to be more abundant following metagenomic versus 16S sequencing, while the reverse was true of *Veillonella, Streptococcus*, and *Collinsella* (**Supplementary Figures 1B and 1C**). Several genera that were rare or absent from 16S data were present in ≥20% of samples based on metagenomic sequencing (e.g., Phocaeicola, Enterocloster, and Coprococcus; **Supplementary Figure 1D**), potentially reflecting discrepancies in reference database alongside removal of 16S-related biases such as primer mismatch. Overall, these findings point to a consistent taxonomic profile across the pipelines, with higher representation of some rarer taxa following metagenomic sequencing.

### Taxonomic differences between Indian and Malawian infants were more substantial than functional differences

Consistent with our previous findings based on 16S data^6^, Malawian infants harboured a more diverse microbiota than Indian infants based on genus-level richness at 6 weeks of age (mean [s.d.] of 55.9 [12.0] vs 45.5 [13.9], respectively; Wilcoxon P value <0.001; **Figure 1A**). This difference in alpha diversity was also evident based on species-level taxonomic profile (149.2 [27.6] vs 123.6 [32.1], respectively; Wilcoxon P value <0.001). Functional profiles yielded low relative abundances of mapped reads in Malawi and India at both pathway level (6.6% [1.0] vs 6.7% [1.1], respectively) and EC level (12.8% [2.1] vs 13.0% [2.0], respectively). Nonetheless, a diverse range of pathways and ECs were assigned in each country. In contrast to the taxonomic profiles, we observed no significant differences in feature richness between Malawian and Indian infants at pathway level (315.6 [44.2] vs 318.3 [51.7], respectively; Wilcoxon P value = 0.145) or EC level (1,444.8 [185.5] vs 1,452.2 [210.6], respectively; Wilcoxon *P* value = 0.396). Significant differences in beta diversity based on unweighted Bray–Curtis distances were evident at genus, species, and EC level (PERMANOVA R^2^ of 3.5%, 5.8%, and 1.2%, respectively; P values <0.05) but not pathway level (R^2^ of 0.5%; *P* = 0.199). Consistent with these findings, Random Forests models were more accurate based on taxonomic profiles than functional profiles (**Figure 1B)**, although all models outperformed the baseline accuracy of 50%.

**Figure 1:**
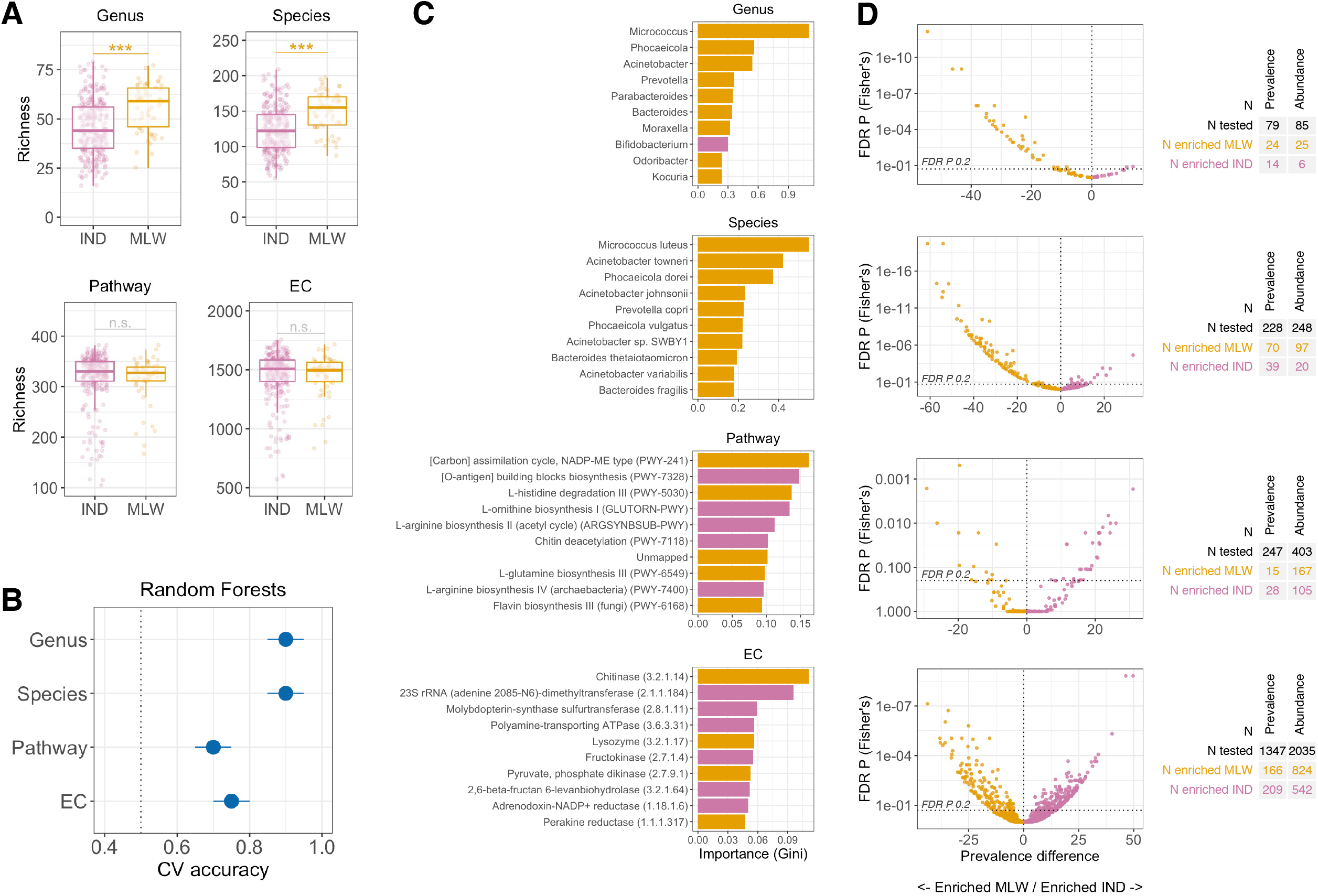
Geographic discrepancies in taxonomic and functional composition of the microbiome. (A) Comparison of feature richness for each data module. Groups were compared by Wilcoxon rank-sum test. (B) Cross-validation accuracy of Random Forests for prediction of country. Median out-of-bag accuracy (proportion correctly assigned) and interquartile range across 20 iterations of 5-fold cross-validation are displayed. A random subset of 50 samples per country was used for each iteration. (C) The 10 most important genera selected by Random Forests for discriminating infants by country. (D) Volcano plots for feature enrichment by country. CV, cross-validation; EC, enzyme commission; FDR, false discovery rate; IND, India; MLW, Malawi; n.s., not significant; *** p <0.0005.

Large numbers of discriminant features were evident across all data modules whether considering their abundance as a continuous variable (via linear regression using MaAsLin2) or presence or absence as a binary variable (via Fisher’s exact test; **Supplementary File 1**). At genus level, 31 features (of 85 tested) were discriminant (FDR P <0.2) based on feature abundance, of which 25 were enriched in Malawi. Findings were similar when considering feature prevalence, wherein 38 genera differed significantly by country, of which 24 were more prevalent in Malawi (**Figure 1C**). The ten top-ranking genera (based on feature importance) selected by Random Forests included nine that were enriched in Malawi (*Microcococcus, Phocaeicola, Acinetobacter*, and *Bacteroides*, among others; Figure 1C left panel) and Bifidobacterium, which was significantly more abundant in India (mean [s.d.] abundance of 59.8% [30.0] and 33.5% [28.6] in India and Malawi, respectively; **Supplementary File 1A**). Findings were consistent at species level, with 117 discriminant features (of 248 tested) identified by MaAsLin2, of which 97 were enriched in Malawi. The top-ranking species selected by Random Forests were consistent with the genus-level selections (*e*.*g. Micrococcus spp*., *Acinetobacter spp*., and *Phoaceicola spp*., all of which were enriched in Malawi; Supplementary File 1B) while offering greater taxonomic resolution of these discriminant taxa.

A total of 272 discriminant pathways (of 403 tested) were identified by MaAsLin2, of which 167 were enriched in Malawi. Among the top-ranking pathways selected by Random Forests, L-glutamine biosynthesis (PWY-6549), L-histidine degradation (PWY-5030), and flavin biosynthesis (PWY-6168) were enriched in Malawi, while O-antigen biosynthesis (PWY-7328), L-ornithine biosynthesis (GLUTORN-PWY), and L-arginine biosynthesis (ARGSYNBSUB-PWY) were enriched in India **(Supplementary File 1C)**, indicative of distinct country-specific metabolic profiles associated with the developing microbiome. EC abundances were the most feature-rich data module. Among 2,035 enzymes tested, 1,366 differed significantly by country based on MaAsLin2, of which 824 were enriched in Malawi. Again, the top-ranking features selected by Random Forests indicated the emergence of distinct country-specific metabolic profiles, with chitinase (3.2.1.14), lysozyme (3.2.1.17), and pyruvate phosphate dikinase (2.7.9.1) among the enzymes enriched in Malawi, with hydroxyethylthiazole kinase (2.7.1.50; involved in thiamine metabolism), acetylornithine transaminase (2.6.1.11; involved in arginine biosynthesis), and alanine transaminase (2.6.1.2; also involved in arginine biosynthesis) among the enzymes enriched in India (**Supplementary File 1D**). In each data module, Random Forests importance scores were highly correlated with MaAsLin2 P values (rho of 0.62, 0.67, 0.31, and 0.30 for genus, species, pathway, and EC data, respectively; P values <0.001), suggesting that features with the most significant abundance discrepancies underpinned the predictive accuracy of these models. Together, these findings demonstrate the developing microbiome of Malawian and Indian infants to be highly distinct in terms of both taxonomic composition and (to a somewhat lesser extent) functional capacity, though suboptimal annotation of functional pathways and ECs may undermine the identification of discriminant features in the latter data modules.

### Breastfeeding status, gut inflammation, and HIV exposure status are associated with taxonomic and functional profile of the developing microbiome

We performed an exploratory analysis of alpha and beta diversity to identify cofactors associated with the taxonomic and functional profile of the developing microbiome in each cohort, analogous to our previous exploratory analysis of 16S data^6^. In Indian infants, 25 demographic and clinical covariates were assessed, including several markers of environmental enteric dysfunction (EED). PERMANOVA of unweighted Bray–Curtis distances revealed microbiome composition to be significantly correlated with breastfeeding status (exclusive vs non-exclusive) across every data module (R^2^ values of 1.9%, 2.4%, 2.4%, and 2.2% for genus, species, pathway, and EC data, respectively; FDR P values <0.2), with exclusive breastfeeding associated with lower feature richness in each instance (**Figure 2** and **Supplementary File 2**). No other covariates were significantly correlated with pathway or EC profile. However, both genus and species composition were significantly associated with mode of delivery, sex, maternal education, and the EED marker α1-antitrypsin.

**Figure 2:**
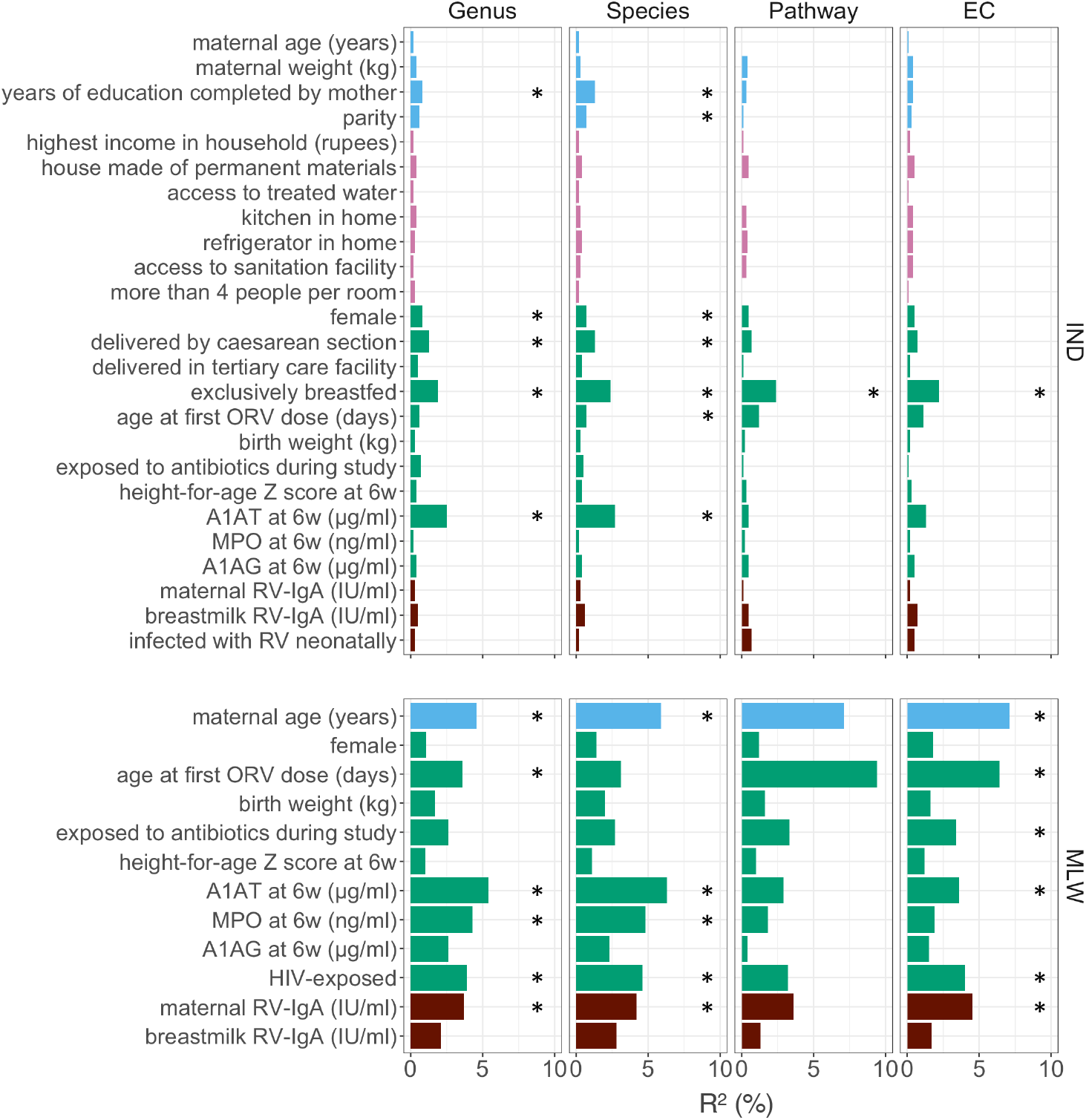
Exploratory analysis of covariates associated with microbiome composition. Factors associated with beta diversity at week of life 6 in Indian and Malawian infants. PERMANOVA was performed using unweighted Bray–Curtis distances for each data module. A1AT, α1-antitrypsin; A1AG, α1 acid glycoprotein; IND, India; MLW, Malawi; MPO, myeloperoxidase; ORV, oral rotavirus vaccine; RV, rotavirus; * FDR p <0.05.

In Malawi, fewer demographic covariates were measured in the cohort and several covariates (e.g., breastfeeding status and delivery mode) lacked sufficient variability for inclusion in this exploratory analysis. Nonetheless, across both taxonomic and functional data modules, microbiome composition was significantly associated with HIV exposure status, markers of EED (including α1-antitrypsin and myeloperoxidase), maternal age, and maternal serum RV-IgA concentration (**Supplementary File 2**). Significant associations with feature richness were generally absent, except for maternal RV-IgA concentration, which was positively correlated with EC richness.

### Taxonomic and functional richness is negatively correlated with ORV seroconversion

Based on 16S profiles, we previously reported genus-level alpha diversity to be significantly higher in ORV non-seroconverters than seroconverters in Malawian infants and Indian infants lacking neonatal rotavirus exposure^6^. We observed the same associations here based on genus- and species-level metagenomic profiles (**Figures 3A** and **3B**), suggesting these trends to be robust to sequencing method and taxonomic resolution. Pathway and EC richness was also negatively correlated with ORV seroconversion (**Figures 3C** and **3D**), although the associations were statistically significant only in Indian infants. In contrast to the genus- and species-level associations, discrepancies in pathway and EC richness were significant in the subset of Indian infants with neonatal rotavirus exposure.

**Figure 3:**
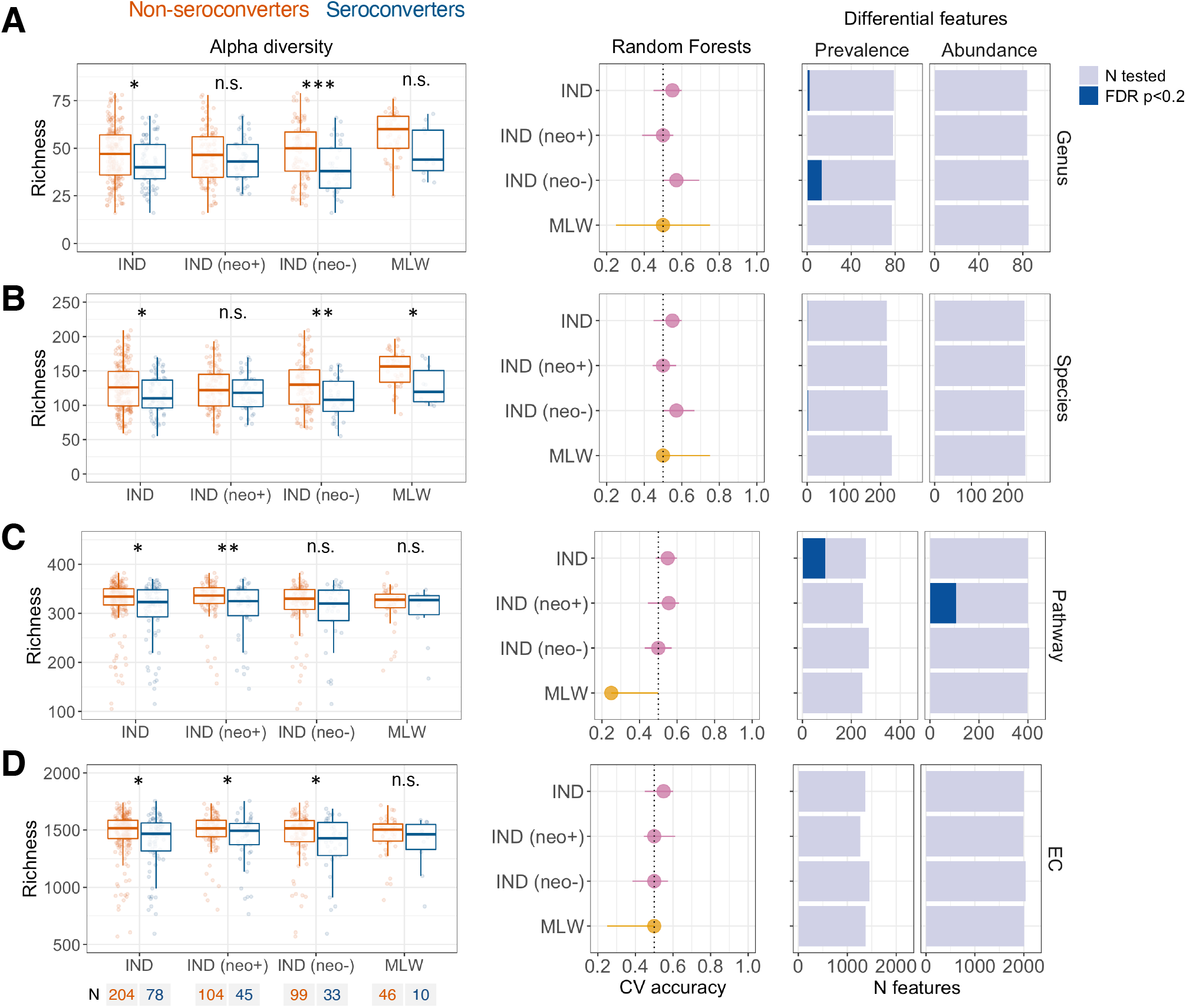
Association between microbiome composition and oral rotavirus vaccine seroconversion. Data are shown for (A) genus, (B), species, (C) pathway, and (D) enzyme commission data. The left panels display comparisons of feature richness by seroconversion. Groups were compared by Wilcoxon rank-sum test. The middle panels display the cross-validation accuracy of Random Forests for prediction of seroconversion. Median out-of-bag accuracy (proportion correctly assigned) and interquartile range across 20 iterations of 5-fold cross-validation are displayed. Each iteration included an equal number of responders and non-responders (50 per group where possible, or else the number in the minority group if this was <50). The right panels display the number of enriched features based on prevalence (Fisher’s exact test) and abundance (MaAsLin2). Analyses for Indian infants are reported for the overall cohort and stratified by neonatal wild-type rotavirus exposure. CV, cross-validation; EC, enzyme commission; FDR, false discovery rate; IND, India; MLW, Malawi; n.s., not significant; neo+, infected with rotavirus neonatally (defined by detection of rotavirus shedding in week of life 1 or baseline seropositivity); neo-, uninfected with rotavirus neonatally; * p <0.05; ** p <0.005; *** p <0.0005.

Random Forests models exhibited poor predictive accuracy for seroconversion across all modules of input data in all cohorts (cross-validation accuracies of <60%; baseline accuracy 50%; **Figures 3A** to **3D**, middle panels). While a small proportion of genera, species, and pathways exhibited differential prevalence or abundance according to seroconversion status (FDR P values <0.2; **Figures 3A** to 3D right panels; **Supplementary File S3**), these were specific to Indian infants and FDR-adjusted P values were typically close to the significance threshold (e.g. of 107 differentially abundant pathways in Indian infants with neonatal rotavirus exposure, none had an FDR-adjusted *P* value of <0.1). We identified no ECs with differential prevalence or abundance according to seroconversion status in any cohort. Together, these findings suggest that individual taxonomic or functional features did not exhibit marked discrepancies according to ORV seroconversion status, which is consistent with our prior findings based on 16S data.

Given the lack of clear discrepancies in feature abundance or prevalence profile, we sought to identify the taxonomic and functional features underpinning the significant differences in richness between seroconverters and non-seroconverters in each cohort. Indeed, while associations for individual features tended not to be statistically significant, there was a clear skew towards features being more prevalent in non-seroconverters than seroconverters (*P* <0.005 for all data modules in all cohorts for test of skew in negative vs positive associations; **Figure 4**). Notably, this skew was highly significant even in comparisons in which all FDR *P* values were above >0.2 (e.g. pathway and EC analyses in Malawian infants).

**Figure 4:**
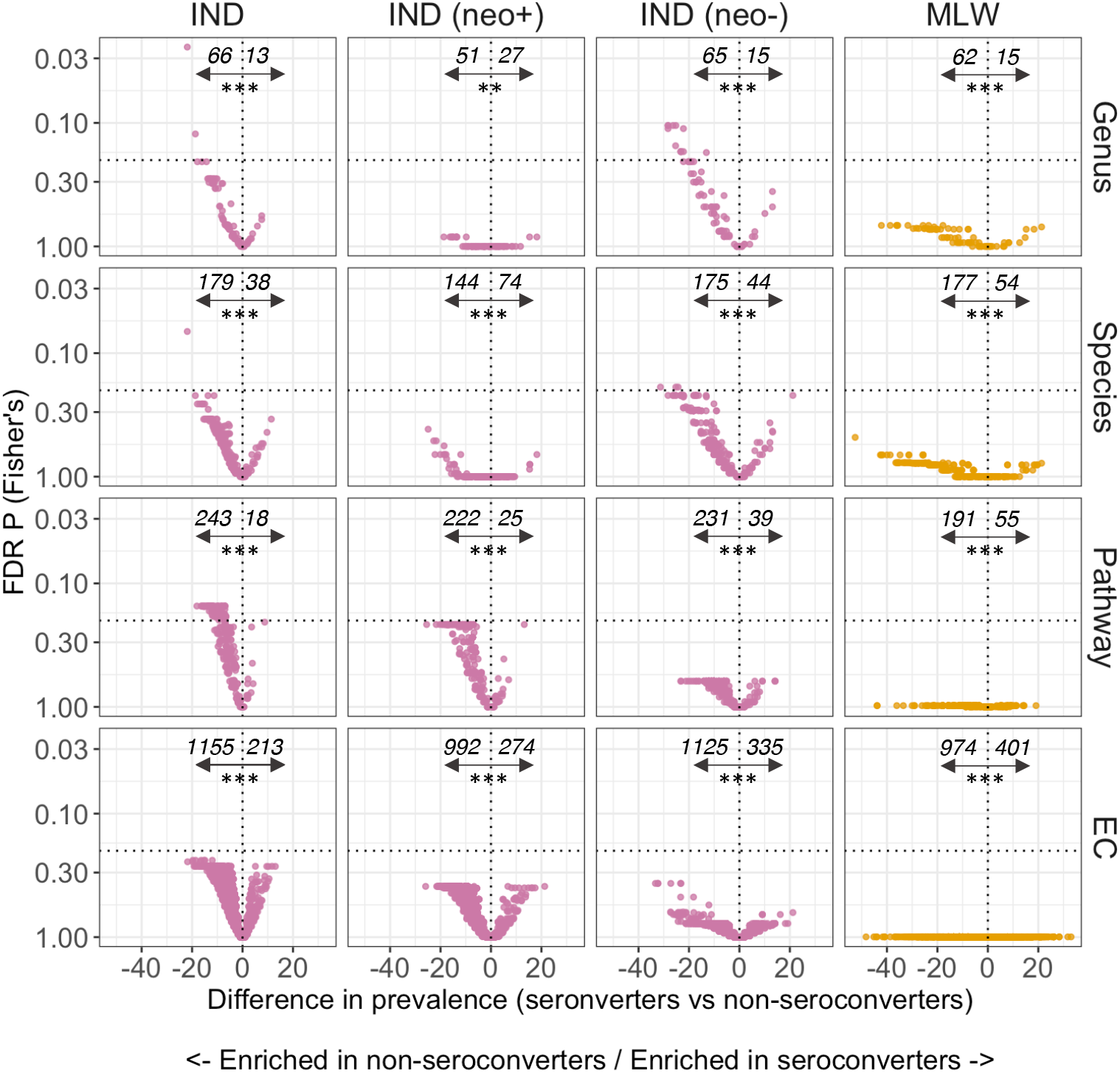
Volcano plots of feature enrichment by oral rotavirus vaccine seroconversion. Italicised numbers display the number of features with a prevalence difference of <0 (left of vertical dotted line) or ≥0 (right of vertical dotted line), highlighting features enriched in non-seroconverters and seroconverters, respectively. The horizontal dotted line indicates an FDR p value of 0.2. The significance tests below the italicised numbers indicate the results of a Wilcoxon rank-sum test assessing skew in the distribution of associations (with a null hypothesis that negative and positive associations occur with equal frequency). Analyses for Indian infants are reported for the overall cohort and stratified by neonatal wild-type rotavirus exposure. EC, enzyme commission; FDR, false discovery rate; IND, India; MLW, Malawi; neo+, infected with rotavirus neonatally (defined by detection of rotavirus shedding in week of life 1 or baseline seropositivity); neo-, uninfected with rotavirus neonatally; sero+, seroconverters; sero-, non-seroconverters; ** p <0.005; *** p <0.0005.

For each data module, we therefore compared the strongest associations according to seroconversion status between Malawian and Indian infants. We assessed the top 10% of association per module (10, 20, 25, and 125 for genus, species, pathway, and EC data, respectively). To enhance comparability across cohorts, we focused on the subset of Indian infants with no neonatal rotavirus exposure. This revealed several notable consistencies between cohorts, as summarised in **Supplementary File S4**. At genus level, 3 of the 10 strongest associations (*Flavonifractor, Odoribacter*, and *Olsenella*) were consistent in India and Malawi – all three genera were more prevalent (by 13–36%) among non-seroconverters than seroconverters in both cohorts. At species level, five of the 20 strongest associations were consistent in India and Malawi – *Intestinimonas butyriciproducens, Flavonifractor plautii, Olsenella* sp. GAM18, *Parabacteroides merdae*, and *Phocaeicola coprophilus*. Again, all were more prevalent (by 22–42%) among non-seroconverters versus seroconverters in both cohorts. At pathway level, four of the 25 strongest associations were consistent in India and Malawi (purine nucleobases degradation I [P164-PWY], peptidoglycan biosynthesis V [beta-lactam resistance; PWY-6470], isopropanol biosynthesis [PWY-6876], and beta-(1,4)-mannan degradation [PWY-7456]). Finally, at EC level, 31 of the 125 strongest associations were consistent across cohorts. These included enzymes associated with adhesion, cell wall degradation, and efflux, among others. The majority of enzymes (27 of 31 [87%]) were more prevalent among non-seroconverters than seroconverters in both cohorts.

Post-vaccination RV-IgA concentration and dose 1 shedding status were considered as alternative measures of ORV response. In contrast to seroconversion status, we documented no significant associations between alpha diversity and ORV response for these endpoints in either country (**Supplementary Figures 2** and **3**). Random Forests models exhibited poor predictive accuracy for these outcomes, and few discriminant features were identified based on prevalence- or abundance-based tests. Despite the lack of significant associations with feature richness, we documented a significant skew towards negative associations in Malawian infants and Indian infants lacking neonatal rotavirus exposure (**Supplementary Figures 4** and **5**; **Supplementary Files 5** and **6**).

For post-vaccination RV-IgA, 1/10, 3/20, 3/25 and 14/125 of the strongest associations were consistent across cohorts at genus, species, pathway and EC level, respectively (**Supplementary File S6)**. Corresponding rates for dose 1 shedding were 1/10, 1/20, 4/25, and 14/125. Notably, these rates do not deviate substantially from the degree of correspondence expected by chance (10%), and there was minimal overlap in the strongest associations selected for different ORV outcomes.

## DISCUSSION

The infant gut microbiome exists in a state of dynamic flux, and host–microbe interactions in this critical developmental window may have long-term ripple effects for immune system development. We used metagenomic sequencing to profile the gut microbiome among Indian and Malawian infants at the time of their first dose of ORV. We observed clear differences between countries in taxonomic and (to a lesser extent) functional composition of the developing microbiome. While the abundances of genera, species, pathways, and enzymes – either individually or collectively – served as poor predictors of ORV response, there was a clear trend towards higher microbiome diversity in ORV non-responders compared with responders. Notably, this trend was consistent across Indian and Malawian infants, and was evident whether considering taxonomic diversity (at genus or species level) or functional capacity (at pathway or enzyme level).

The geographic differences in taxonomic composition reported here (e.g. enrichment of *Bifidobacterium, Klebsiella*, and *Enterococcus* in India vs enrichment of *Escherichia, Streptococcus*, and *Bacteroide*s in Malawi) are generally consistent with our prior study based on 16S sequencing^6^. However, several discriminant genera were detected only via metagenomic sequencing (e.g. enrichment of *Micrococcus, Phocaeicola*, and *Moraxella* in Malawi), likely reflecting the greater sequencing depth obtained in the current study and the reduction in biases (e.g. primer mismatch) when applying metagenomic as opposed to 16S sequencing.

The proportion of reads assigned at pathway or enzyme level was low (7% and 13%, respectively), consistent with prior studies applying the HUMAnN pipeline to samples from LMICs^15^. This highlights the significant under-representation of reference genes necessary for precise functional annotation of LMIC samples in UniRef90. This is an ongoing, pervasive issue in microbiome analysis^16,17^, underpinned by a systemic lack of representation in human microbiome databases. Recent work highlighting this issue demonstrated that 0.7% of all microbiome data in the Sequence Read Archive, DNA Data Bank of Japan and European Nucleotide Archive is derived from India and Malawi^18^. As such, the levels of read assignment observed in our metagenomic dataset is unsurprising. Attempts to improve equity in genomic research capacity, such as H3Africa^19^, have begun to address this issue. This caveat notwithstanding, there was greater consistency between countries when viewing the microbiome with a functional rather than taxonomic lens, as typified by the lower predictive accuracy of Random Forests models and the strong overlap in abundant pathways and enzymes between Indian and Malawian samples.

Several previous studies have explored the link between infant oral vaccine response and microbiome composition. A metagenomics study in Zimbabwe documented no significant associations between ORV seroconversion and either the composition or function of the microbiome^19^, which is consistent with the lack of discriminatory features based on genus, species, pathway, or enzyme abundance in the present study. Among infants in Ghana, Rotarix immunogenicity was positively correlated with the relative abundance of 16S-based sequence variants assigned to the genus *Streptococcus* and the family Enterobacteriaceae^20^, partially recapitulating earlier findings from the same cohort^21^. We did not identify similar taxonomic discrepancies according to seroconversion status here, although several taxa (e.g. *Flavonifractor plautii* and *Parabacteroides merdae*) were among the top-ranking associations in both India and Malawi and may merit further causal exploration if validated elsewhere. Likewise, several pathways (e.g. beta-(1,4)-mannan degradation and peptidoglycan biosynthesis V) and ECs (e.g. Xaa-Xaa-Pro tripeptidyl-peptidase and blood group B branched chain alpha-1,3-galactosidase were enriched in non-seroconverters versus seroconverters in both countries, highlighting functional components of the developing microbiome that may be pertinent to ORV response. As the body of literature exploring the link between the developing microbiome and ORV response expands, robust taxonomic or functional associations across independent cohorts remain elusive, and those identified lack causal validation. Although this does not preclude the possibility of microbiota-directed therapeutic to enhance ORV immunogenicity, the selection of potential candidates for such an intervention have thus far failed to emerge from observational studies in affected populations.

A key finding in the present study was the higher diversity of the microbiome among non-seroconverters compared with seroconverters, which was apparent in both Malawian and Indian infants. We previously observed the same association in both cohorts using 16S-based methods^6^. The present study therefore serves as a validation of the robustness of this association, both to sequencing method (16S vs metagenomics) and annotation type (taxonomic vs functional). Notably, other studies of ORV in LMICs have not reported a significant association between alpha diversity and seroconversion^21,22^, although we previously documented a negative correlation between microbiota diversity and oral poliovirus vaccine shedding among Indian infants aged 6–11 months^23^. A causal link between greater microbiome diversity and impaired oral vaccine response is possible and merits further consideration. For example, differences in innate immune responses between children from different regions can be partly recapitulated via transfer of stool microbiomes to germ-free mice^24^, and microbiome diversity may be among the key factors shaping innate immune development in the intestinal epithelium.

Our findings sit somewhat at odds with a growing body of literature highlighting the beneficial effects of a taxonomically diverse microbiome^26,27^. It is worth noting that much of the existing microbiome literature is biased towards conditions affecting adult populations in high-income countries, whereas we focused on risk factors for impaired oral vaccine response in LMICs. While we do not discount the broad health benefits of a diverse microbiome, it may be that diverse microbial stimulation in early life is protective against some conditions (e.g. autoimmune and inflammatory disorders) while contributing to others, such as the blunting of oral vaccine responsiveness in LMICs. Moving forward, the search for markers of a ‘healthy microbiome’ should be tuned to population- and age-specific morbidity indicators, and may not yield consistent answers as a result.

The strengths of this study lie in the use of standardised methods across diverse LMIC cohorts, randomised order to reduce batch effects, and validation of prior findings using novel sequencing and bioinformatic methods, lending robustness to the reported trends. However, there are several limitations to this work, including sub-optimal annotation, which left much of the functional landscape uncharted. As reference databases become more representative, it may be that future analyses of our metagenomic data yield additional insights. We also experienced recruitment and sample collection challenges in Malawi, leaving this cohort underpowered (see previous publication^6^ for details). Moreover, our pipeline and methods were tailored to the bacterial microbiome, so may miss an important role of other branches of the developing microbiome, such as the virome or eukaryome.

Overall, we applied metagenomic sequencing to validate and extend findings of a multi-centre cohort study exploring risk factors for impaired ORV response. We found the developing microbiome to be more diverse – both in terms of its taxonomic composition and functional capacity – among non-seroconverters compared with seroconverters in India and Malawi. By confirming this trend across multiple LMIC cohorts, our findings suggest that higher microbiome diversity in early life may act as marker for impaired oral vaccine response.

## Supporting information

Supplementary File 1 - country

Supplementary File 2 - cofactors

Supplementary File 3 - seroconversion

Supplementary File 4 - top features

Supplementary File 5 - shedding

Supplementary File 6 - IgA

## Data Availability

Outputs from our analyses are available within the supplementary material of this manuscript. Sequencing data are currently undergoing submission to the European Nucleotide Archive. Scripts used for analysis will be made available on GitHub.

## Acknowledgements

We dedicate this paper to our friend and colleague Dr Sudhir Babji. The Malawi site of the RoVI study was funded by the UK Medical Research Council and the UK Department for International Development (Newton Fund MR/N006259/1). The site in India was funded by the Government of India’s Department of Biotechnology. Metagenomic sequencing was supported by the IMmunising PRegnant women and INfants neTwork (IMPRINT), funded by the GCRF Networks in Vaccines Research and Development, which was co-funded by the UK MRC and UK BBSRC. This UK funded award is part of the EDCTP2 programme supported by the European Union. Library preparation and sequencing was performed at the Centre for Genomic Research at the University of Liverpool. E.C.-O. and A.C.D. are affiliated to the National Institute for Health Research (NIHR) Health Protection Research Unit in Gastrointestinal Infections at University of Liverpool, in partnership with the UK Health Security Agency (UKHSA), in collaboration with University of Warwick. E.C.-O. and A.C.D. are based at The University of Liverpool. The views expressed are those of the author(s) and not necessarily those of the NIHR, the Department of Health and Social Care or the UK Health Security Agency.

## Figure legends

**Supplementary Figure 1:**
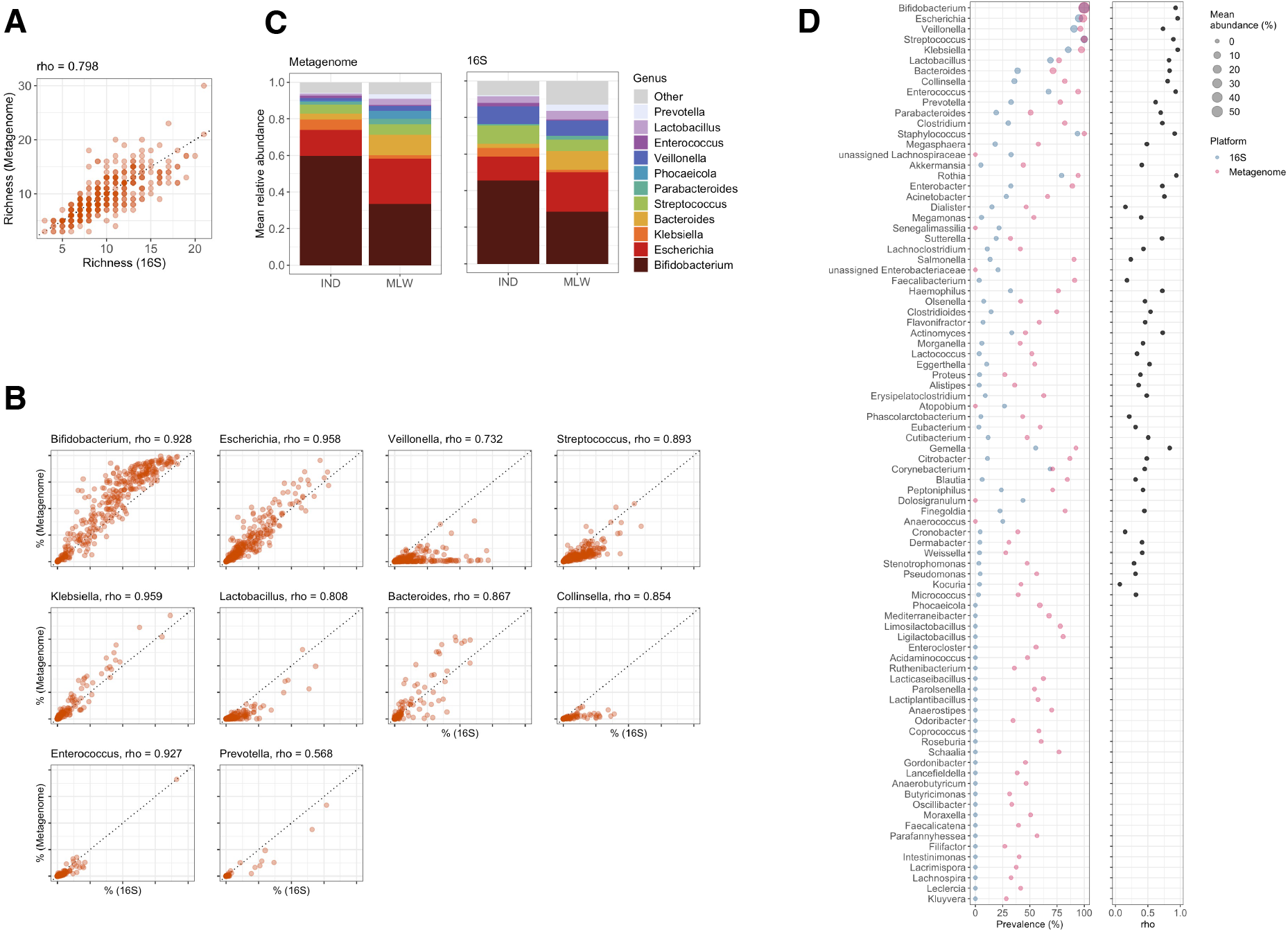
Comparison of 16S and metagenomic sequencing profiles. (A) Comparison of genus-level richness estimates. Correlation was assessed via Spearman’s rank correlation coefficient (rho). (B) Comparison of genus relative abundances for 10 post common genera in 16S data. Mean genus abundances by country. Genera with a mean abundance of at least 1% in either country based on either metagenomic or 16S profile are displayed. (D) Prevalence, abundance, and correlation profiles for genera present in at least 20% of samples based on either metagenomic or 16S profile. Correlation was assessed via Spearman’s rank correlation coefficient (rho) for genera detected by both platforms.

**Supplementary Figure 2:**
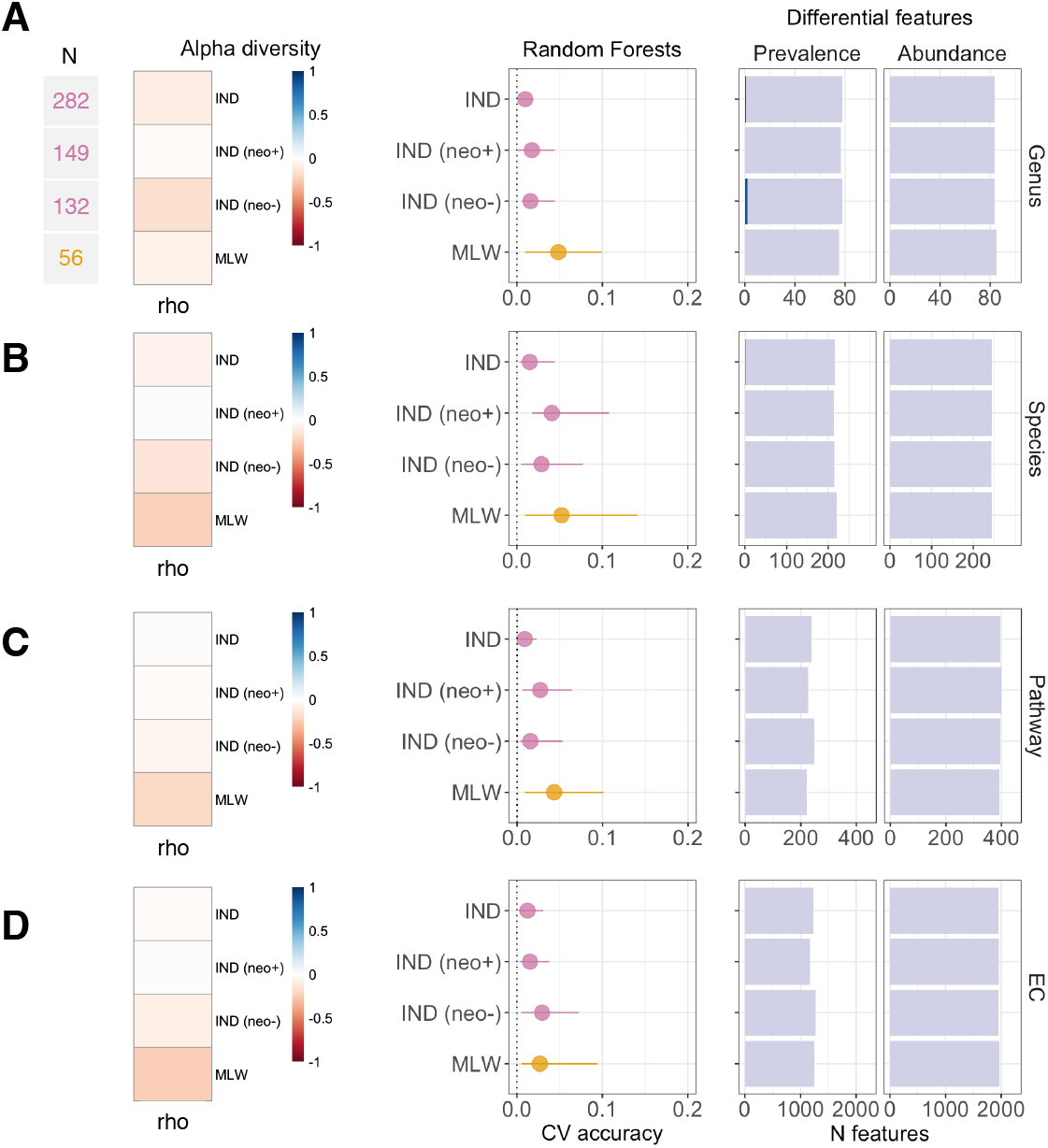
Association between microbiome composition and rotavirus-specific IgA. Data are shown for (A) genus, (B), species, (C) pathway, and (D) enzyme commission data. The left panels display correlations between feature richness and post-vaccination RV-IgA concentration status, as determined via Spearman’s rank correlation coefficient (rho) with two-sided hypothesis testing. The middle panels display the cross-validation accuracy of Random Forests for prediction of post-vaccination RV-IgA. Median out-of-bag R_2_ and interquartile range are displayed for predicted vs observed RV-IgA across 20 iterations of 5-fold cross-validation. The right panels display the number of enriched features based on prevalence (Wilcoxon rank-sum test for RV-IgA concentration in individuals with versus without the feature in question) and abundance (MaAsLin2). Analyses for Indian infants are reported for the overall cohort and stratified by neonatal wild-type rotavirus exposure. CV, cross-validation; EC, enzyme commission; FDR, false discovery rate; IND, India; MLW, Malawi; n.s., not significant; neo+, infected with rotavirus neonatally (defined by detection of rotavirus shedding in week of life 1 or baseline seropositivity); neo-, uninfected with rotavirus neonatally.

**Supplementary Figure 3:**
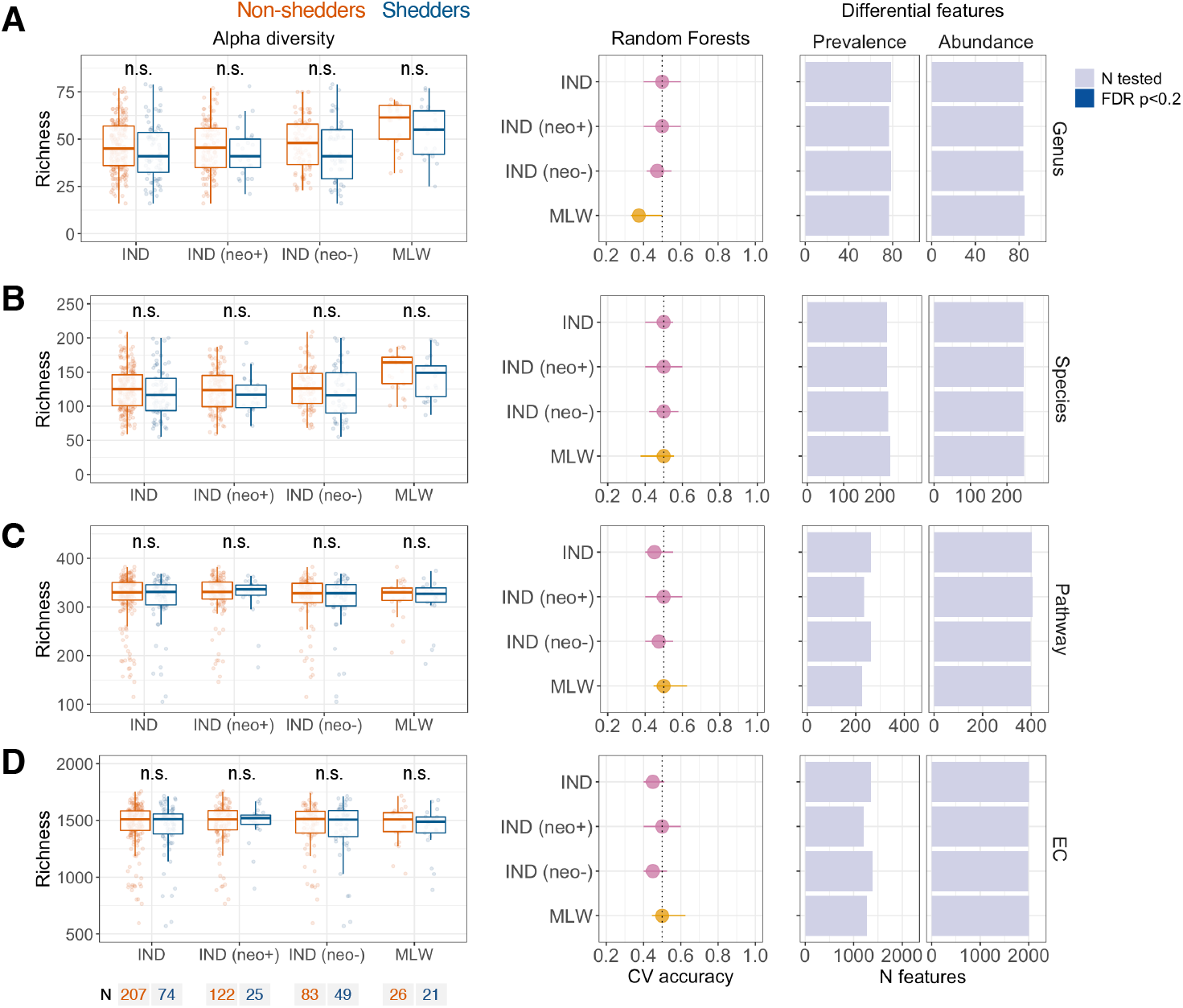
Association between microbiome composition and dose 1 oral rotavirus vaccine shedding. Data are shown for (A) genus, (B), species, (C) pathway, and (D) enzyme commission data. The left panels display comparisons feature richness by shedding status. Groups were compared by Wilcoxon rank-sum test. The middle panels display the cross-validation accuracy of Random Forests for prediction of shedding status. Median out-of-bag accuracy (proportion correctly assigned) and interquartile range across 20 iterations of 5-fold cross-validation are displayed. Each iteration included an equal number of responders and non-responders (50 per group where possible, or else the number in the minority group if this was <50). The right panels display the number of enriched features based on prevalence (Fisher’s exact test) and abundance (MaAsLin2). Analyses for Indian infants are reported for the overall cohort and stratified by neonatal wild-type rotavirus exposure. CV, cross-validation; EC, enzyme commission; FDR, false discovery rate; IND, India; MLW, Malawi; n.s., not significant; neo+, infected with rotavirus neonatally (defined by detection of rotavirus shedding in week of life 1 or baseline seropositivity); neo-, uninfected with rotavirus neonatally.

**Supplementary Figure 4:**
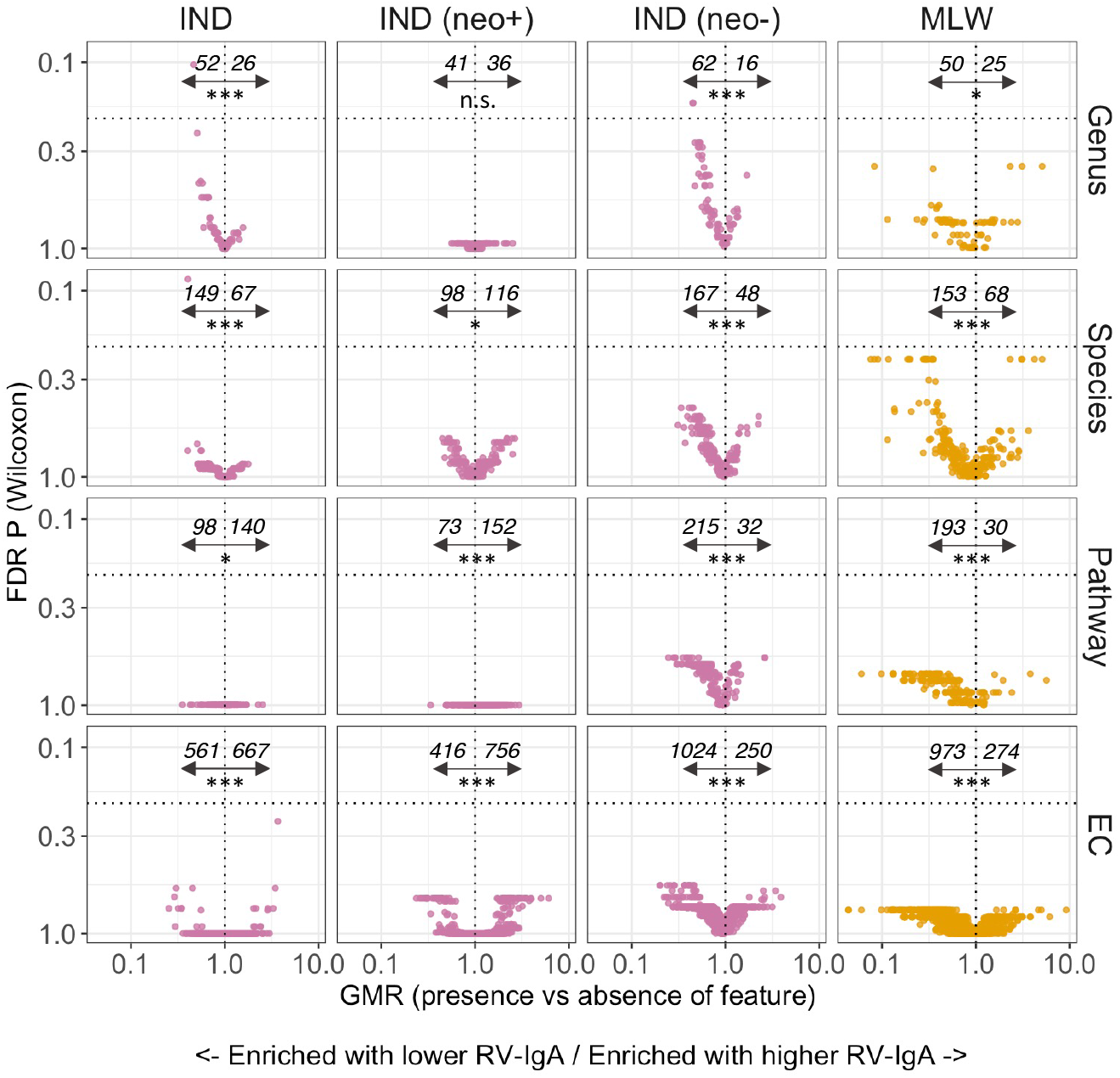
Volcano plots of geometric mean rotavirus-specific IgA ratios in relation to feature presence or absence. Italicised numbers display the number of features with a prevalence difference of <1 (left of vertical dotted line) or ≥1 (right of vertical dotted line), highlighting features associated with lower and higher RV-IgA levels, respectively. The horizontal dotted line indicates an FDR p value of 0.2. The significance tests below the italicised numbers indicate the results of a Wilcoxon rank-sum test assessing skew in the distribution of associations (with a null hypothesis that negative and positive associations occur with equal frequency). Analyses for Indian infants are reported for the overall cohort and stratified by neonatal wild-type rotavirus exposure. EC, enzyme commission; FDR, false discovery rate; GMR, geometric mean ratio; IND, India; MLW, Malawi; n.s., not significant; neo+, infected with rotavirus neonatally (defined by detection of rotavirus shedding in week of life 1 or baseline seropositivity); neo-, uninfected with rotavirus neonatally; * p <0.05; *** p <0.0005.

**Supplementary Figure 5:**
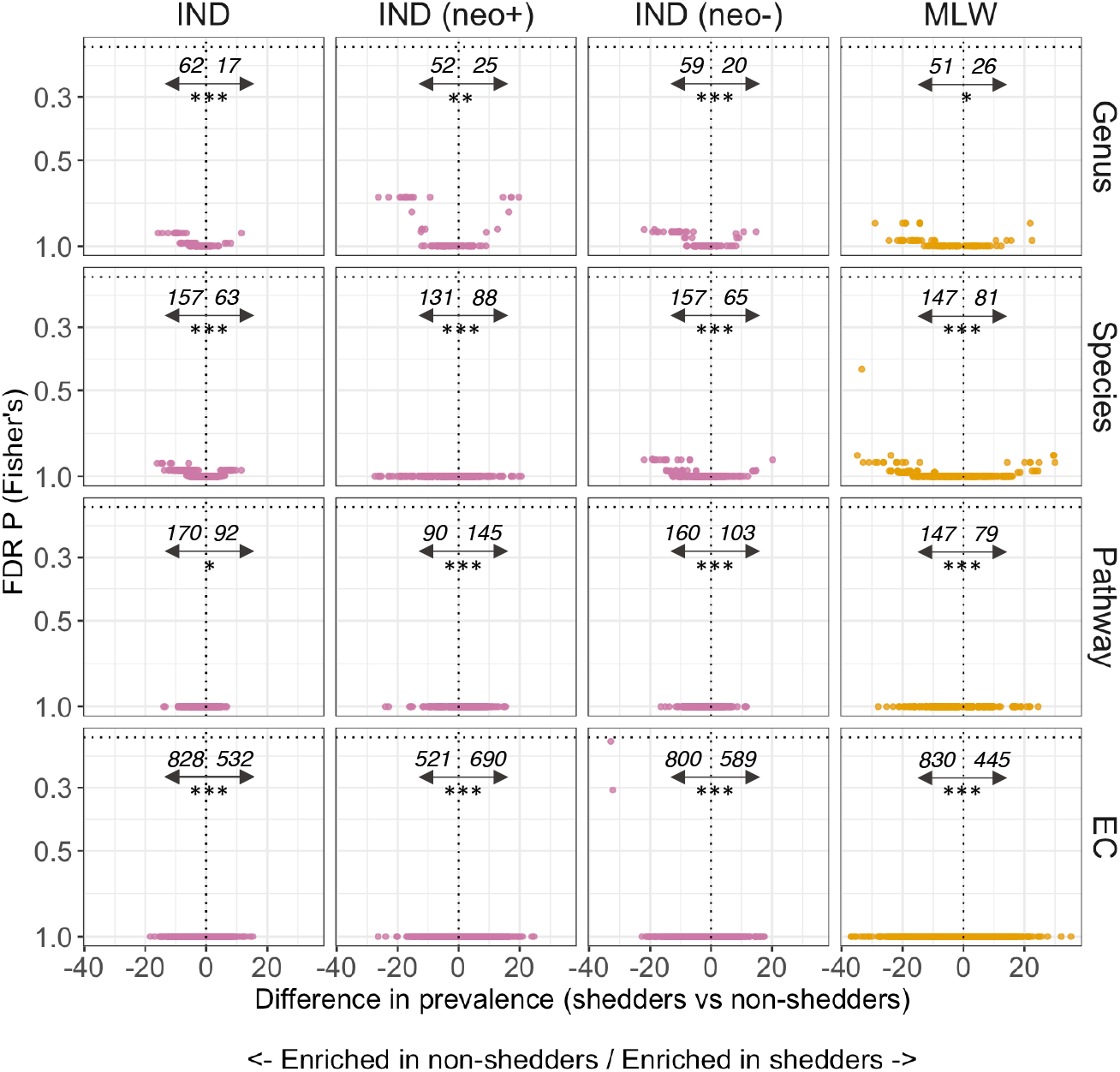
Volcano plots of feature prevalence in relation to dose 1 rotavirus vaccine shedding. Italicised numbers display the number of features with a prevalence difference of <0 (left of vertical dotted line) or ≥0 (right of vertical dotted line), highlighting features enriched in shedders and shedders, respectively. The horizontal dotted line indicates an FDR p value of 0.2. 2. The significance tests below the italicised numbers indicate the results of a Wilcoxon rank-sum test assessing skew in the distribution of associations (with a null hypothesis that negative and positive associations occur with equal frequency). Analyses for Indian infants are reported for the overall cohort and stratified by neonatal wild-type rotavirus exposure. EC, enzyme commission; FDR, false discovery rate; IND, India; MLW, Malawi; neo+, infected with rotavirus neonatally (defined by detection of rotavirus shedding in week of life 1 or baseline seropositivity); neo-, uninfected with rotavirus neonatally; sero+, seroconverters; sero-, non-seroconverters; ** p <0.005; *** p <0.0005.

